# ARE GEOGRAPHIC FACTORS ASSOCIATED WITH POORER OUTCOMES IN PATIENTS DIAGNOSED WITH COVID-19?

**DOI:** 10.1101/2021.01.25.21250404

**Authors:** Rosa Magallón-Botaya, Bárbara Oliván-Blázquez, Karen Lizzette Ramirez-Cervantes, Fatima Mendez-Lopez de la Mazanara, Marc Casajuana-Closas, Eva Esteban-Andrés

**Affiliations:** Health Research Institute of Aragon (IIS Aragon). Spain; Department of Psychology and Sociology. University of Zaragoza. Zaragoza. Spain; Department of Medicine. University of Zaragoza. Zaragoza. Spain; University of Rey Juan Carlos, Madrid. Research group in Management in the Bleeding Patient. Spain; University Autonomous of Barcelona. Spain; Institute of Research Hospital La Paz (IdlPaz); Spanish Association aginst Cancer. Madrid; Institut of Research of Primary Care Jordi Gol (IDIAP Jordi Gol), Barcelona, Spain; Primary Care Prevention and Health Promotion Network (redIAPP). Institute of Health Carlos III. Spain

## Abstract

**Background:** The prognosis of patients with COVID-19, with older age and comorbidities, is associated with a more severe course and higher fatality rates but no analysis has yet included factors related to the geographical area/municipality in which the affected patients live. So the objective of this study is to analyse the prognosis of patients with COVID-19 in terms of sex, age, comorbidities, and geographic variables.

**Methods:** A retrospective cohort of 6286 patients diagnosed with COVID-19 was analysed, considering demographic data, previous comorbidities and geographic variables. The main study variables were hospital admission, Intensive Care Unit (ICU) admission and death due to worsening symptoms; and the secondary variables were sex, age, comorbidities and geographic variables (size of the area of residence, distance to the hospital and the driving time to the hospital). A comparison analysis and a multivariate Cox model were performed.

**Results:** The multivariate Cox model showed that women had a better prognosis in any type of analysed prognosis. Most of the comorbidities studied were related to a poorer prognosis except for dementia, which is related to lower admissions and higher mortality. Suburban areas were associated with greater mortality and with less hospital or ICU admission. Distance to the hospital was also associated with hospital admission.

**Conclusions:** Factors such as type of municipality and distance to hospital act as social health determinants. This fact must be taken account in order to stablish specifics prevention measures and treatment protocols.

## BACKGROUND

As of the 5^th^ of January 2021, more than 83 million cases of the novel coronavirus disease 2019 (COVID-19) have been detected worldwide, and more than 1.8 million people have died from the disease ^1^. Spain has become one of the countries most affected by the disease, and one of the countries with the highest COVID-19 testing rates^2,3,4^.

The response of National Health Service (NHS), especially that of Primary Health Care (PHC), has been crucial for containing COVID-19 ^5^. Spain has included maintaining essential services by cancelling or postponing nonurgent activities and elective surgeries. Moreover, barriers to accessing usual care were mitigated by enhancing phone helplines, online assistance and emergency call centres ^6^. Furthermore, the PHC services had to be reorganised according to their rural, urban or suburban characteristics, and home care and telemedicine were reinforced ^3^. The first medical contact for COVID-19 cases is commonly through a PHC team, which determines the severity of symptoms, manages the follow-up of mild cases and organises the hospital referrals for moderate-severe cases ^3^. However, the urban-rural gap could have affected the way in which the current health crisis was confronted ^7^. In 2019, for instance, people over the age of 65 years represented 28.5% of the population of rural communities (≤2000 inhabitants) compared with 19.7% and 18.3% of suburban and urban communities, respectively ^8^. In addition, 42% of older adults from rural areas considered that distance posed a major difficulty in accessing a PHC centre, which, along with a greater dependency for performing activities of daily living for this group and a lack of public transportation, could have caused an uneven impact of COVID-19 ^9^.

The Spanish NHS is decentralised; each of the 17 autonomous communities is responsible for the territorial administration of Primary and Hospital Health Services ^10^. There are 13,163 PHC centres in Spain ^11^; however, due to asymmetric population density, the centres are not uniformly distributed, with basic health areas the smallest units of health care, each covering 2000–10,000 inhabitants. As observed in other territories ^12,13^, the current statistics show that the incidence of COVID-19 has been unequal across Spain ^14^. For instance, recent Catalan studies have shown that the spread of the disease was slower in rural territories than in urban areas, with 6.5% higher in basic health areas with twice the population density ^15^. However, the introduction of an infected individual into small but dense groups, can negatively impact the spread of the disease, as observed in several rural communities where a high incidence rate was related to the presence of a COVID-19-positive individual at highly attended funerals.

In countries such as India, 25% of COVID-19-related deaths have occurred in rural and suburban districts ^12^. In the United States, the COVID-19 incidence and mortality rates of small and nonmetropolitan cities have been comparable to those of large cities.^13^

An increasing number of articles have been addressing the prognosis of patients with COVID-19, showing that older age and comorbidities, were associated with a more severe course and higher fatality rates among individuals hospitalised for COVID-19 ^16–18^ To our knowledge, no analysis has related factors related to the geographical area/municipality in which the affected patients live and their geographic accessibility to a referral hospital.

In countries like Spain with large rural areas and low population densities, the effects of spatial disparities on disease outcomes are still unknown. The objective of this study was therefore to analyse the prognosis (hospital admission, Intensive Care Unit [ICU] admission and death) of patients with COVID-19 in terms of sex, age, comorbidities, and geographics variables.

## METHOD

We conducted a retrospective cohort study that considered demographic data, previous comorbidities and geographics variables of 6286 patients diagnosed with COVID-19 in Aragon (Spain) from the beginning of the current pandemic to the 30^th^ of June 2020.

The inclusion criteria were an age older than 18 years and a diagnostic code of “coronavirus infection”. There were no exclusion criteria.

Due to the universal nature of the NHS, the data obtained are considered representative of practically 100% of the population who met the criteria for inclusion.

Aragón has an area of 47,719 km^2^ and a population density of 28.20 inhabitants per km^2^, with a higher proportion of older adults than younger ones. The older adults are more concentrated in rural areas, while the cities have younger populations. Zaragoza, the capital, contains half of the community’s population, with only 13 municipalities exceeding 10,000 inhabitants. Rural nuclei (with fewer than 2000 inhabitants) represent 86% of the municipalities, where only 16.8% of the population lives.

The population pyramid graph for Aragon ^14^ shows a contracting structure. The main age group is 30–49 years (active population), which has been augmented by a significant increase in the immigrant population since 2000. However, population older than 65 years has grown significantly since the beginning of the century, a common feature in all developed regions. The origins of this situation include a substantial reduction in fertility rates, and an increase in life expectancy at birth. This region’s geographical and demographic characteristics make it comparable to many other regions of inland Spain and other European and American countries with high levels of ageing, geographical dispersion and depopulation.

The pyramid shows low crude birth rates due to the small number of initial cohorts (aged 0–4 years) and the low crude death rates.

The primary study variables were: Hospital admission, ICU admission and Death due to COVID-19.

The Secondary Variables were the following:

- Sociodemographic data: sex and age
- Previous medical history: previous Cardiovascular Diseases (CVD): chronic heart disease, heart failure, cerebrovascular disease, Hypertension, Dyslipidaemia, previous Chronic Respiratory Diseases, Chronic Renal Diseases, Chronic Liver diseases, Chronic Neurological disorders, chronic haematological diseases (leukaemia, lymphoma, myeloma), cancer/neoplasia, HIV and other immunodeficiencies, Obesity, Diabetes, Dermatological diseases, Rheumatological diseases, mental disorders and dementia. The diseases were classified according to the International Classification of Diseases 11th Revision ^19^.
- Geographic variables: The size of the area of residence (rural areas were defied as municipalities with a population of fewer than 2000 inhabitants, suburban areas were defined as towns with 2000-10,000 inhabitants, and urban areas were defined as having more than 10,000 inhabitants), municipalities with nursing home care (yes/no), distance (Km) from the residence to the hospital and the driving time (minutes) from the residence to the hospital.

For Statistical Analysis, we extracted all data on the demographics, clinical treatments and outcomes from the Electronic Medical Records. We described the quantitative variables using robust statistics, such as mean and interquartile range (IQR) and analysed the qualitative data based on their frequency distribution. For the comparison of quantitative data between groups, we employed a Mann–Whitney nonparametric U test. For the comparison of qualitative variables between the good/poor prognosis groups, we used the chi-squared test. We performed the survival estimates with the Kaplan–Meier method, comparing the survival curves according to the prognosis groups; the Wilcoxon test did not reach the median survival due to the survival curves. Multivariate analysis was performed by a Cox regression with the forward conditional method, introducing as independent variables the poorest prognosis factor. The results of the multivariate model are presented as a hazard ratio (HR; 95% CI).

The statistical analysis was performed using STATA/SE V16.0, and p-values <0.05 were considered statistically significant.

All procedures contributing to this study complied with the ethical standards of the Helsinki Declaration of 1975, (revised in 2008). The study protocol was approved by the Clinical Research Ethics Committee of Aragón. (PI20/262). Clinical data were used in a nonidentifiable format.

## RESULTS

Of the 6286 patients studied, 2738 (43.56%) were men, 4440 (71.72%) lived in urban areas, and the median age was 61 years (IQR 61-82). Table 1 lists the demographic characteristics and the prevalence of previous diseases. The patients were significantly older in the poorest prognosis groups, regardless of whether the patients were in the hospital admission, ICU admission or death groups. The female sex was associated with a better prognosis (Table 1). With a considerably lower ICU admission rate, than among men (28.7% vs 62.2%) and a lower final mortality rate (48.5% vs 57.9%) and hospital admission rate (47.6% vs 62.2%) than the men.

**Table 1:** Sociodemographic data and previous comorbidities according to different prognoses

|  |  | Hospital admission |  | P | ICU admission |  | P | Death |  | P |
| --- | --- | --- | --- | --- | --- | --- | --- | --- | --- | --- |
|  |  | No | Yes |  | No | Yes |  | No | Yes |  |
| n |  | 3797<br>(60.40%) | 2489<br>(39.60%) |  | 6034<br>(95.99%) | 252<br>(4.01%) |  | 5313<br>(84.52%) | 973 (15.48%) |  |
| Age* |  | 52 (39-72) | 75 (61-86) | < 0.001 | 45 (60-83) | 67 (59-74) | 0.002 | 56 (43-74) | 86 (79-92) | < 0.001 |
| Sex |  |  |  | < 0.001 |  |  | < 0.001 |  |  | < 0.001 |
|  | Male | 1434<br>(37.77%) | 1304<br>(52.39%) |  | 2557<br>(42.38%) | 181<br>(71.83%) |  | 2237<br>(42.10%) | 501 (51.49%) |  |
|  | Female | 2363<br>(62.23%) | 1185<br>(47.61%) |  | 3477<br>(57.62%) | 71<br>(28.71%) |  | 3076<br>(57.90%) | 472 (48.51%) |  |
| Municipality classification |  |  |  | 0.002 |  |  | 0.009 |  |  | < 0.001 |
|  | Rural | 430<br>(11.52%) | 302<br>(12.24%) |  | 696 (11.69%) | 36<br>(14.63%) |  | 622 (11.88%) | 110 (11.40%) |  |
|  | Suburban | 559<br>(17.93%) | 358<br>(14.51%) |  | 1003<br>(16.85%) | 24 (9.76%) |  | 803 (15.34%) | 224 (23.21%) |  |
|  | Urban | 2633<br>(70.55%) | 1807<br>(73.25%) |  | 4254<br>(71.46%) | 186<br>(75.61%) |  | 3809<br>(72.77%) | 631 (65.39%) |  |
| Municipality - hospital distance, km* |  | 0 (0-27.55) | 0 (0-18.70) | < 0.001 | 0 (0-25.20) | 0 (0-17.70) | 0.136 | 0 (0-25.20) | 0 (0-27.70) | 0.018 |
| Drive time from municipality to hospital, min* |  | 0 (0-23) | 0 (0-19) | < 0.001 | 0 (0-23) | 0 (0-17) | 0.127 | 0 (0-22) | 0 (0-25) | 0.015 |

There was a striking correlation with the size of the patients’ area of residence. Patients living in urban areas had poorer prognoses in terms of hospital and ICU admissions. However, this tendency did not occur with respect to mortality, given that 65.46% of the patients who died lived in urban areas compared with 72.87% of those who survived (p < .001). The prevalence rate of patients with COVID-19 in each of the 3 analysed areas shows a high prevalence in the suburban zone, with 48 cases per 10,000 inhabitants in the urban zones, 55 cases per 10,000 inhabitants in the suburban zones, and 35 cases per 10,000 inhabitants in the rural zones. The analysed distance to the hospitals through the median distance and time of arrival to the hospital (Table 1) also proved to be significant.

In terms of previous comorbidity (Table 2), the poorer prognoses were associated with a higher prevalence of any of the diseases studied, except for HIV and other immunodeficiency diseases.

**Table 2:**
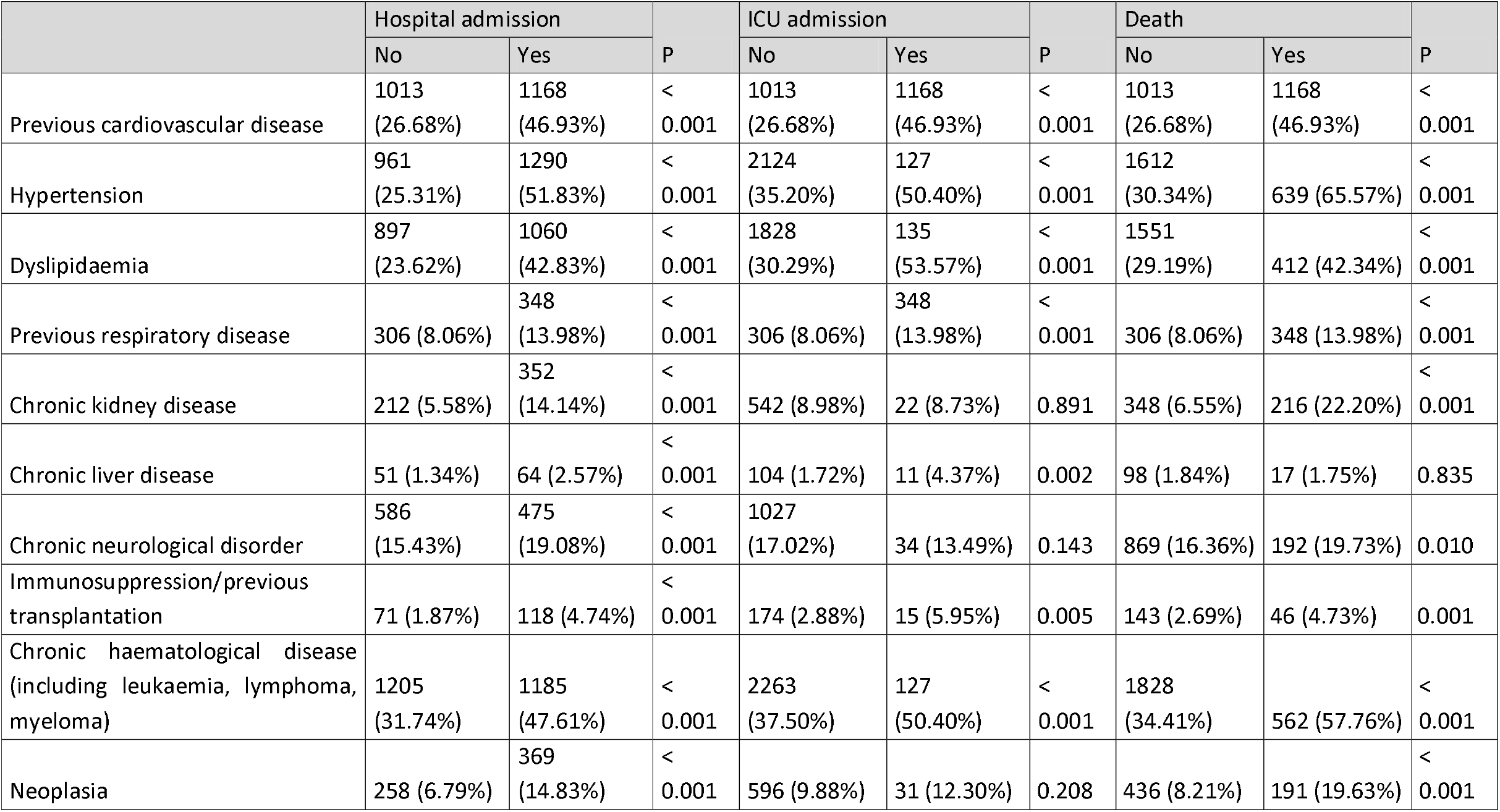
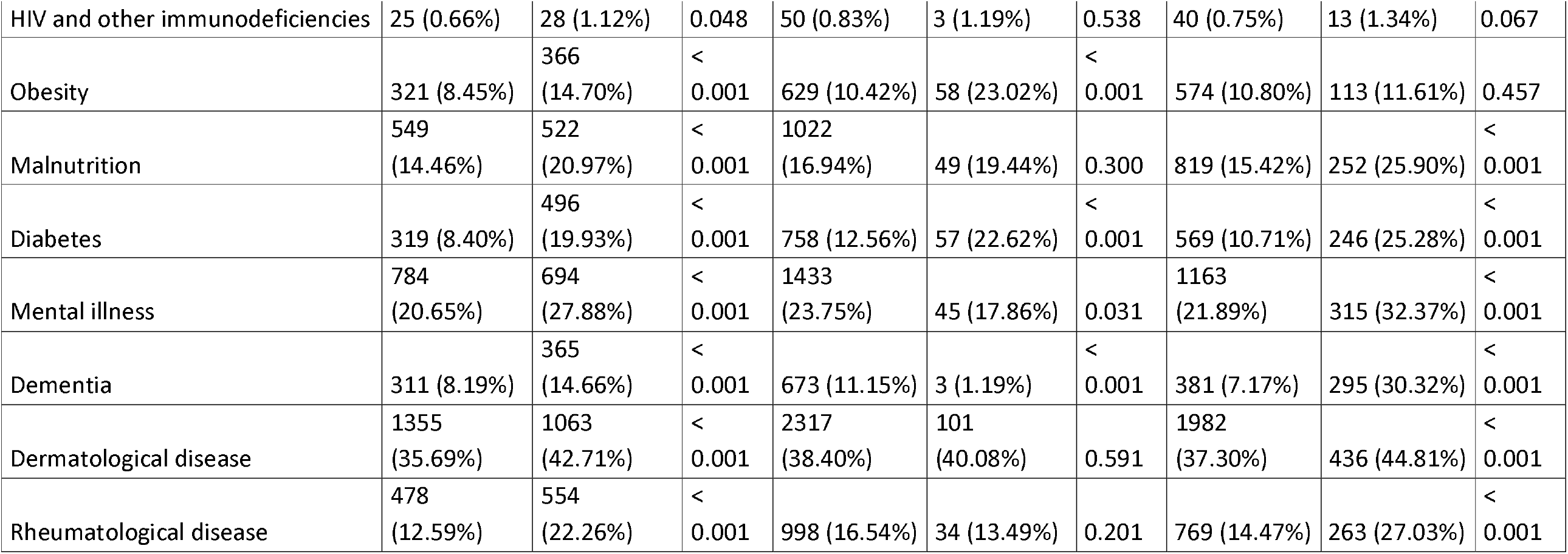
Previous comorbidities

|  | Hospital admission |  |  | ICU admission |  |  | Death |  |  |
| --- | --- | --- | --- | --- | --- | --- | --- | --- | --- |
|  | No | Yes | P | No | Yes | P | No | Yes | P |
| Previous cardiovascular disease | 1013<br>(26.68%) | 1168<br>(46.93%) | <<br>0.001 | 1013<br>(26.68%) | 1168<br>(46.93%) | <<br>0.001 | 1013<br>(26.68%) | 1168<br>(46.93%) | <<br>0.001 |
| Hypertension | 961<br>(25.31%) | 1290<br>(51.83%) | <<br>0.001 | 2124<br>(35.20%) | 127<br>(50.40%) | <<br>0.001 | 1612<br>(30.34%) | 639 (65.57%) | <<br>0.001 |
| Dyslipidaemia | 897<br>(23.62%) | 1060<br>(42.83%) | <<br>0.001 | 1828<br>(30.29%) | 135<br>(53.57%) | <<br>0.001 | 1551<br>(29.19%) | 412 (42.34%) | <<br>0.001 |
| Previous respiratory disease | 306 (8.06%) | 348<br>(13.98%) | <<br>0.001 | 306 (8.06%) | 348<br>(13.98%) | <<br>0.001 | 306 (8.06%) | 348 (13.98%) | <<br>0.001 |
| Chronic kidney disease | 212 (5.58%) | 352<br>(14.14%) | <<br>0.001 | 542 (8.98%) | 22 (8.73%) | 0.891 | 348 (6.55%) | 216 (22.20%) | <<br>0.001 |
| Chronic liver disease | 51 (1.34%) | 64 (2.57%) | <<br>0.001 | 104 (1.72%) | 11 (4.37%) | 0.002 | 98 (1.84%) | 17 (1.75%) | 0.835 |
| Chronic neurological disorder | 586<br>(15.43%) | 475<br>(19.08%) | <<br>0.001 | 1027<br>(17.02%) | 34 (13.49%) | 0.143 | 869 (16.36%) | 192 (19.73%) | 0.010 |
| Immunosuppression/previous transplantation | 71 (1.87%) | 118 (4.74%) | <<br>0.001 | 174 (2.88%) | 15 (5.95%) | 0.005 | 143 (2.69%) | 46 (4.73%) | 0.001 |
| Chronic haematological disease (including leukaemia, lymphoma, myeloma) | 1205<br>(31.74%) | 1185<br>(47.61%) | <<br>0.001 | 2263<br>(37.50%) | 127<br>(50.40%) | <<br>0.001 | 1828<br>(34.41%) | 562 (57.76%) | <<br>0.001 |
| Neoplasia | 258 (6.79%) | 369<br>(14.83%) | <<br>0.001 | 596 (9.88%) | 31 (12.30%) | 0.208 | 436 (8.21%) | 191 (19.63%) | <<br>0.001 |
| HIV and other immunodeficiencies | 25 (0.66%) | 28 (1.12%) | 0.048 | 50 (0.83%) | 3 (1.19%) | 0.538 | 40 (0.75%) | 13 (1.34%) | 0.067 |
| Obesity | 321 (8.45%) | 366 (14.70%) | < 0.001 | 629 (10.42%) | 58 (23.02%) | < 0.001 | 574 (10.80%) | 113 (11.61%) | 0.457 |
| Malnutrition | 549 (14.46%) | 522 (20.97%) | < 0.001 | 1022 (16.94%) | 49 (19.44%) | 0.300 | 819 (15.42%) | 252 (25.90%) | < 0.001 |
| Diabetes | 319 (8.40%) | 496 (19.93%) | < 0.001 | 758 (12.56%) | 57 (22.62%) | < 0.001 | 569 (10.71%) | 246 (25.28%) | < 0.001 |
| Mental illness | 784 (20.65%) | 694 (27.88%) | < 0.001 | 1433 (23.75%) | 45 (17.86%) | 0.031 | 1163 (21.89%) | 315 (32.37%) | < 0.001 |
| Dementia | 311 (8.19%) | 365 (14.66%) | < 0.001 | 673 (11.15%) | 3 (1.19%) | < 0.001 | 381 (7.17%) | 295 (30.32%) | < 0.001 |
| Dermatological disease | 1355 (35.69%) | 1063 (42.71%) | < 0.001 | 2317 (38.40%) | 101 (40.08%) | 0.591 | 1982 (37.30%) | 436 (44.81%) | < 0.001 |
| Rheumatological disease | 478 (12.59%) | 554 (22.26%) | < 0.001 | 998 (16.54%) | 34 (13.49%) | 0.201 | 769 (14.47%) | 263 (27.03%) | < 0.001 |

In the case of ICU admissions, neither CKD (p = .891) nor chronic neurological disease (p = .143) were statistically significant. In the case of mortality, chronic liver disease was the only comorbidity that had a similar distribution among patients who died and those who did not (p = .835).

Figures 1-3 show the patients’ characteristics according to prognosis, with the survival curves according to the size of the residents’ area and the p-value based on the log-rank test. In the Kaplan–Meier curves, we observed that the poorest prognosis was related to suburban areas where fewer people were admitted to hospital than in the other municipalities; nevertheless, the COVID-19-related mortality in these areas was higher than in the other areas studied.

**Figure 1:**
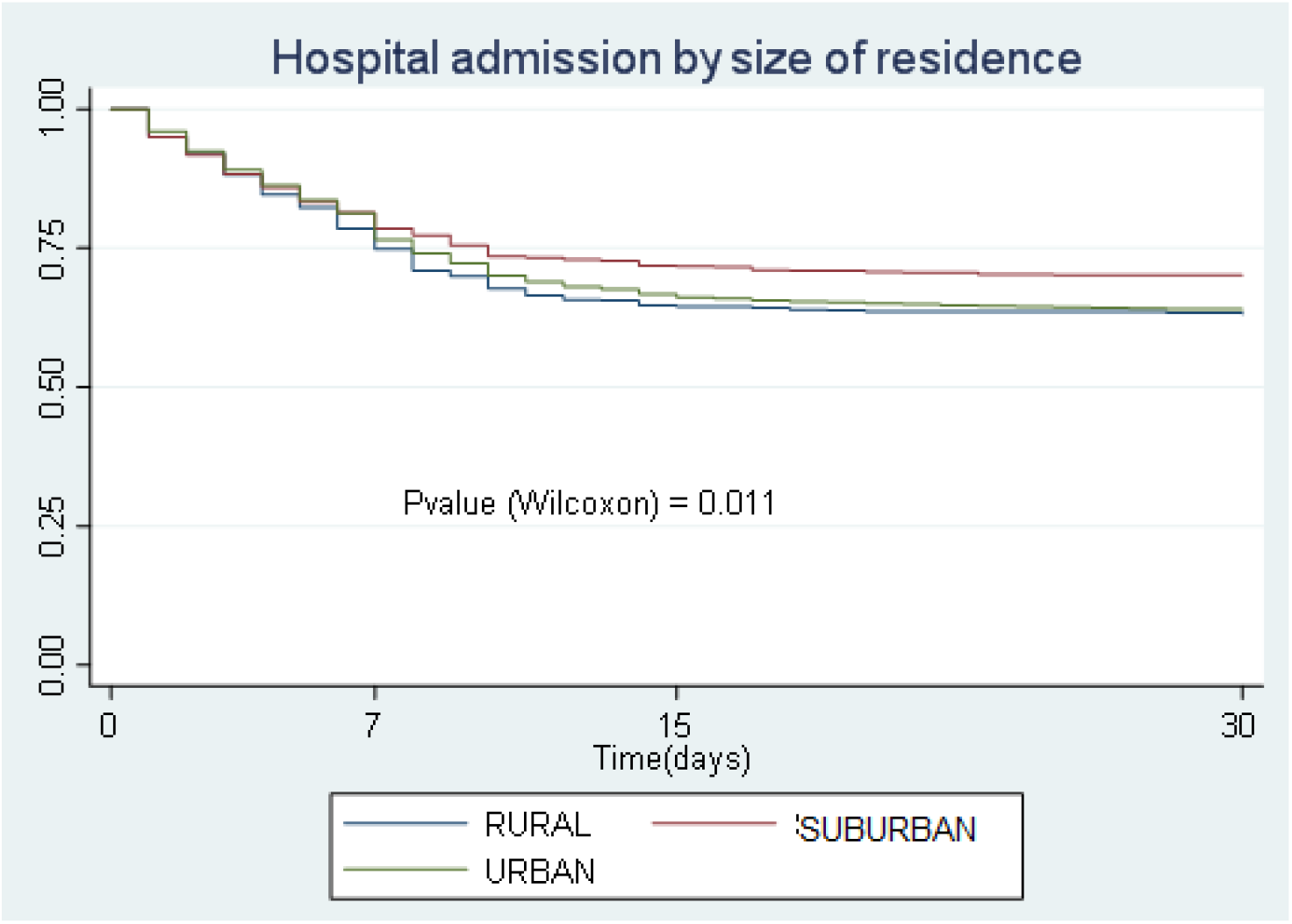
Hospital admission by size of residence, Kaplan–Meier survival.

**Figure 2:**
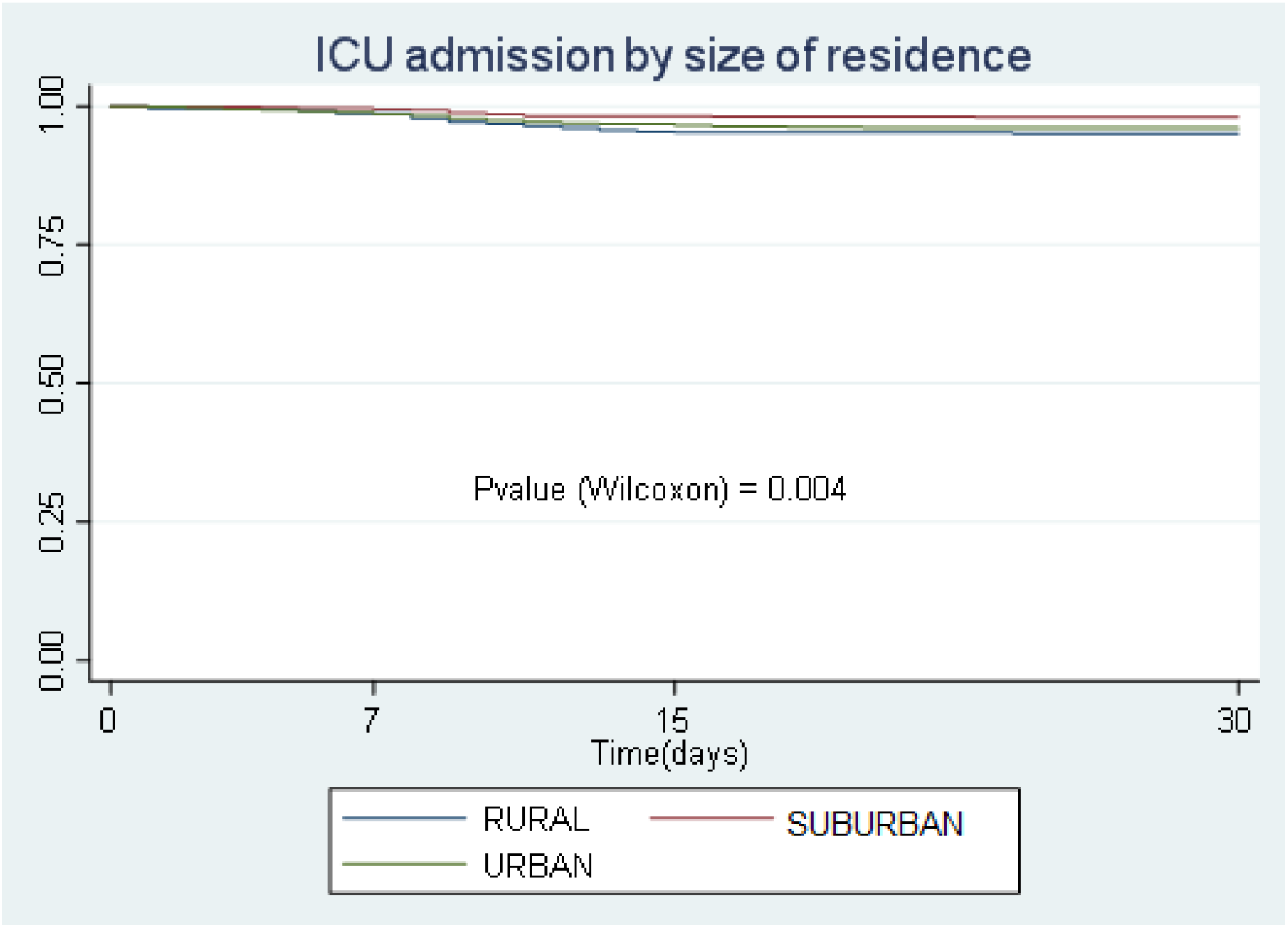
ICU admission by size of residence, Kaplan-Meier survival.

**Figure 3:**
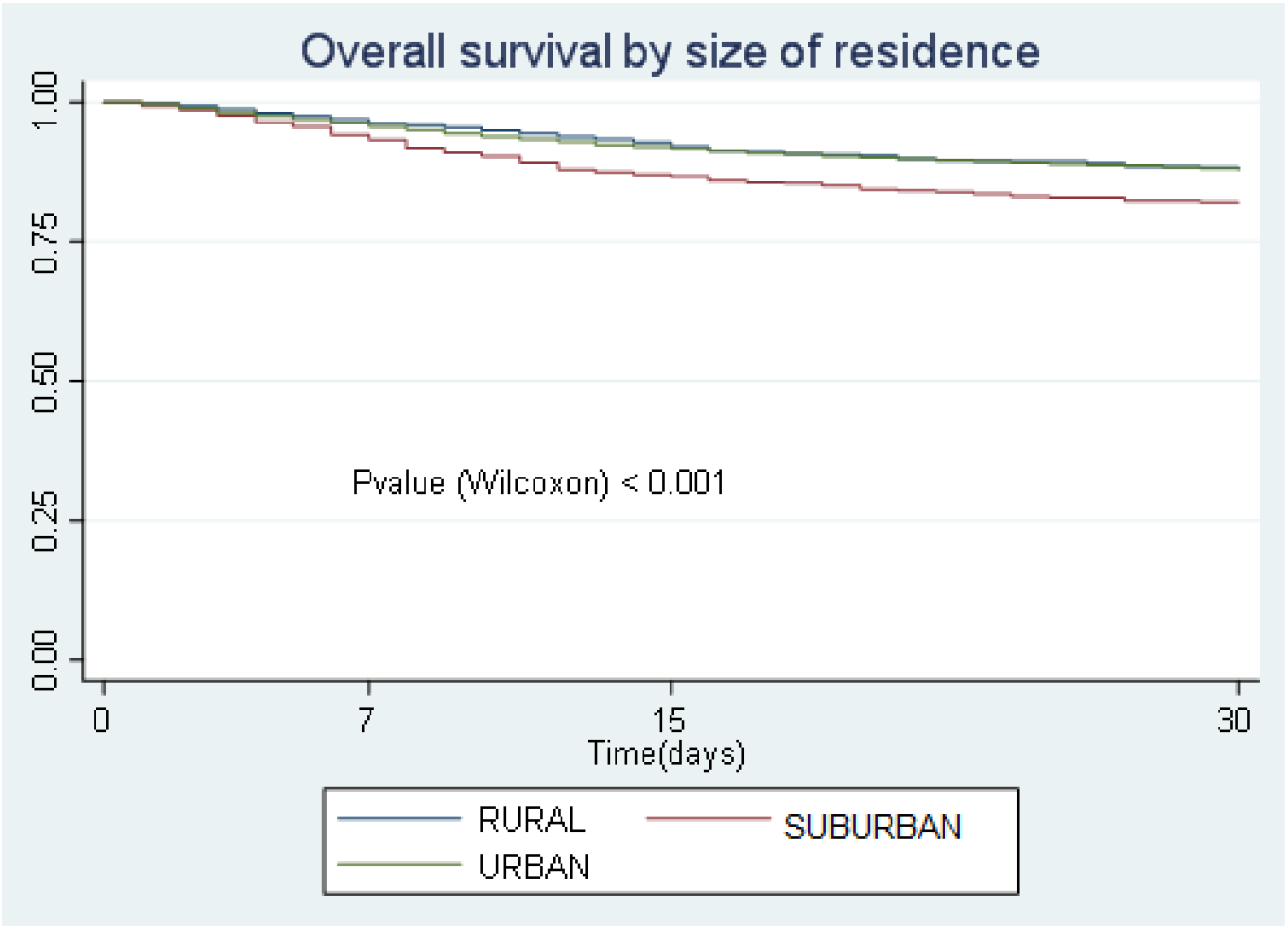
Overall survival by size of residence, Kaplan–Meier survival.

Table 3 shows the multivariate Cox model for each prognosis. As previously noted, women had a better prognosis than men in any type of analysed prognosis. We can also observe similar behaviour in terms of prognosis in the urban and rural areas. Nevertheless, suburban areas were associated with greater mortality and with less hospital or ICU hospitalisation. Distance from the hospital was also associated with hospital admission such that the closer the patients’ residence was to the hospital, the greater the likelihood that they would be admitted.

**Table 3:**
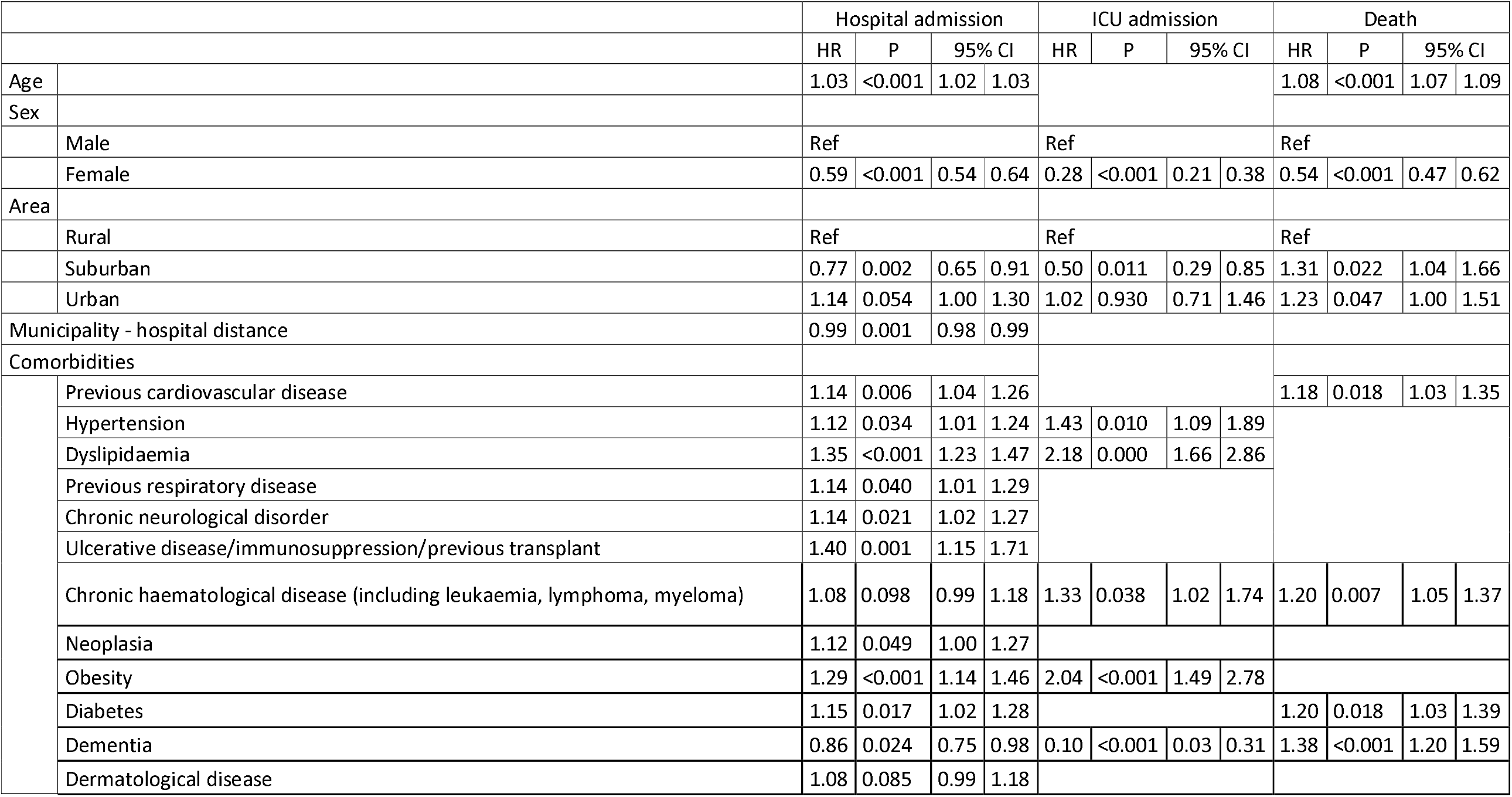
Multivariate Cox models

In terms of the prognostic model of hospital admission, most of the comorbidities studied were related to a poorer prognosis except for dementia (HR 0.86; 95% CI 0.75-0.98), which acts as a protective factor for admissions but is still related to higher mortality.

In the ICU admission model, there were comorbidities such as CVD, neurological disease and previous respiratory disease that disappeared from the final model. Only hypertension (HR 1.43; 95% CI 1.09-1.89), dyslipidaemia (HR 2.18; 95% CI 1.66-2.86), chronic haematological disease (HR 1.33; 95% CI 1.02-1.74) and obesity (HR 2.04; 95% CI 1.49-2.78) were associated with a poorer prognosis in terms of ICU admission.

Lastly, the comorbidities associated with mortality were CVD (HR 1.18; 95% CI 1.03-1.35), previous haematological disease (HR 1.20; 95% CI 1.05-1.37), diabetes (HR 1.20; 95% CI 1.03-1.39) and dementia (HR 1.38; 95% CI 1.20-1.59) (Table 2).

## DISCUSSION

The results of this study show how factors such as type of municipality and distance to hospital can act as social health determinants and establish the outcomes of patients with COVID-19. This study concurs with relevant studies in the literature on the prognosis of patients with COVID-19 while showing that geographical factors are relevant to the prognosis.

One of our relevant results, is that suburban areas were associated with increased mortality and with lower hospital and ICU admission. The reduced accessibility of suburban areas to hospitals compared with urban zones could be a causal factor in this higher mortality; nevertheless, this is not the case in more rural areas, which have shown the same associations as urban areas, despite the fact that, in Spain, individuals older than 65 years accounted for 28.5% of the population of rural communities ^8^. It is probably that small towns, which are typically further away from urban cities, are inhabited by an older population with greater social isolation and less need to travel for work reasons. So, it is highly likely that the older population has been particularly cautious in the face of the pandemic.

Mixed results can be found for the type of municipality. A study conducted in India showed that 1 in 4 COVID-19-related deaths occurred in rural and suburban districts ^12^; however, the incidence and mortality rates in small and nonmetropolitan cities has been comparable to those of large cities in the US ^13^. In our study, 11.64% of the sample lived in rural areas with similar hospitalisation rates than urban areas. In a recent systematic review, the household and area-level social determinants of multimorbidity were analysed ^20^ ; findings from the rural areas were inconsistent and insufficiently studied. In the same review, distance from the hospital was associated with hospitalisation, such that the closer the patient lived to the hospital (urban areas), the greater the likelihood that they would be hospitalised.^20^ In an analysis of avoidable hospitalisations in chronic diseases, the patients’ place of residence explained only 33% of the variation in hospital admission.^21^

Geographic differences in COVID-19 case, deaths and cumulative incidences likely reflect a combination of epidemiologic and population-level factors, including the timing of the start of the pandemic; population density; age distribution and prevalence of underlying medical conditions; timing and extent of community mitigation measures; diagnostic testing capacity; and public health practices ^22^.

Other variables in our study, age and sex, were significant factors in explaining the prognosis of the patients diagnosed in PHC, which agrees with most previous studies that have stated that male and older patients (≥50 years) are at higher risk of greater severity and death ^23–25^. Older adults had greater initial comorbidities, more severe symptoms and are more likely to experience multiorgan involvement ^24^, whereas the sex disparity in the outcomes of patients with COVID-19 could be explained by the fact that men are more likely than women to experience severe forms of infection, and have higher mortality rates and a higher prevalence of the main risk factors of COVID-19 ^26^. Other explanations include the mechanisms of viral infection, the immune response, and the development of hyperinflammation and systemic complications, particularly thromboembolism. Women therefore have a more favourable disease course than men, regardless of age range, although the rate of severe acute respiratory syndrome coronavirus 2 (SARS-CoV-2) infection appears to be similar for both sexes ^25^. Our results reinforce these findings, showing a major difference in ICU admissions by sex, probably due to the lower severity of the disease in women, making ICU admission unnecessary. Women’s greater longevity also means that their deaths occur more frequently in non-hospital settings.

The presence of CVD, diabetes, dementia, hypertension, dyslipidaemia and obesity were relevant to the outcomes of our 6286 patients with COVID-19. Other studies have shown the influence of morbidities in COVID-19. A meta-analysis with 55 studies and 10,014 patients^23^ showed that the presence of at least 1 comorbidity such as hypertension, diabetes, CVD, respiratory disease, CKD, etc. significantly increased the severity of infection. Other studies and meta-analyses have obtained similar results ^17,27–30^.

Hypertension is associated with a nearly 2.5-fold increased risk of severe COVID-19 (OR 2.49; 95% CI 1.98-3.12; I2 = 24%) and a similarly significant higher mortality risk (OR 2.42; 95% CI 1.51-3.90; I2 = 0%)^28^. CVD are also associated with an increased risk of poor outcomes in patients with COVID-19 ^18,29^. Diabetes in patients with COVID-19 is associated with a 2-fold increase in mortality and COVID-19 severity compared with patients without diabetes ^27^; however, this negative impact might not be related to hyperglycaemia per se but rather to the comorbidities associated, in the context of metabolic syndrome, as well as CVD and CKD, which are common and severe complications of chronic hyperglycaemia ^17^.

In our study, severity of COVID-19 appears to increase with increasing body mass index (BMI). A study analysing 45,650 participants from 30 studies with obesity revealed increased ORs of severe COVID-19 associated with higher BMI, ^33^ may be associated to a vitamin D deficiency, hinders immunity and causes mechanical lung compression^32^. Other potential pathophysiological mechanisms are the chronic proinflammatory state, the excessive oxidative stress response and the impaired immunity that is commonly reported in obesity ^31^. Furthermore, pre-existing comorbidities increase these complications ^32^.

Although the patients with dementia in our study had a higher mortality rate, the disease acted as a protective factor for hospital admission, which could be because health systems might be more reluctant to hospitalise patients with dementia and COVID-19, especially in cases of saturated health systems. Patients with preexisting neurological disorders, can develop exacerbation of neurological symptoms and severe COVID-19 ^34,35^. A high percentage of elderly patients with dementia admitted to ICU or isolated medical departments experience neurological and neuropsychiatric symptoms and worsening of their condition ^35^.

The main strengths of this study are its complex model, which includes geographic variables dealing with the comorbidities and prognosis of patients with COVID-19. Our findings can be extrapolated to other countries with similar population pyramid and geographic dispersion.

The current pandemic, like other previous pandemics, is occurring in the context of social and health inequalities that need to be resolved ^37^. This study has sought to contribute knowledge to the current pandemic by analysing the comorbidities associated with a large sample of patients with COVID-19, relating them to health determinants such as age, sex, geographical accessibility to health services and place of residence.

In future studies, data should be compared with those of other countries, especially to provide greater knowledge on the behaviour of health systems and methods for adjusting the use of these systems to this new disease, reducing as much as possible the clinical and epidemiologic variability and exploring the place of residence as a social determinant.

## Data Availability

Aragon Health Service
Institute of Research IIS-Aragon

## Notes

### Competing Interest Statement

The authors have declared no competing interest.

### Funding Statement

Financial support for the conduct of the research was obtained from the Health Institute Carlos III (ISCIII) (grants number COV20_00634 and COV20_00519), and from de Institute of Research IIS-Aragon

### Author Declarations

The study protocol was approved by the Clinical Research Ethics Committee of Aragon: PI20/262

